# Tear antibodies to SARS-CoV-2: implications for transmission

**DOI:** 10.1101/2021.08.02.21261479

**Authors:** Kevin J. Selva, Samantha K. Davis, Ebene R. Haycroft, Wen Shi Lee, Ester Lopez, Arnold Reynaldi, Miles P. Davenport, Helen E. Kent, Jennifer A. Juno, Amy W. Chung, Stephen J. Kent

## Abstract

**Objectives:** SARS-CoV-2 can be transmitted by aerosols and the ocular surface may be an important route of transmission. Little is known about protective antibody responses to SARS-CoV-2 in tears after infection or vaccination. We analysed SARS-CoV-2 specific IgG and IgA responses in human tears after either COVID-19 infection or vaccination.

**Methods:** We recruited 16 subjects with COVID-19 infection an average of 7 months previously and 15 subjects before and 2 weeks after Comirnaty (Pfizer-BioNtech) vaccination. Plasma, saliva and basal tears were collected. Pre-pandemic plasma, saliva and basal tears from 11 individuals were included as healthy controls. Antibody responses to 5 SARS-CoV-2 antigens were measured via multiplex.

**Results:** IgG antibodies to Spike and Nucleoprotein were detected in tears, saliva and plasma from subjects with prior SARS-CoV-2 infection in comparison to uninfected controls. While RBD-specific antibodies were detected in plasma, minimal RBD-specific antibodies were detected in tears and saliva. In contrast, high levels of IgG antibodies to Spike and RBD, but not Nucleoprotein, were induced in tears, saliva and plasma of subjects receiving 2 doses of the Comirnaty vaccine. Increased levels of IgA1 and IgA2 antibodies to SARS-CoV-2 antigens were detected in plasma following infection or vaccination, but were unchanged in tears and saliva.

**Conclusion:** Both infection and vaccination induce SARS-CoV-2-specific IgG antibodies in tears. RBD-specific IgG antibodies in tears were induced by vaccination but were not present 7 months post-infection. This suggests neutralising antibodies may be low in the tears late following infection.

## Text

SARS-CoV-2, the cause of the COVID-19 pandemic, is commonly acquired via airborne transmission. Observational studies show that the wearing of spectacles is associated with reduced SARS-CoV-2 acquisition, suggesting SARS-CoV-2 may be acquired through the ocular surface [1, 2]. Similarly, eye protection initiatives have been associated with reduced health-care associated outbreaks of COVID-19 [3]. As a result of these findings, eye protection is widely recommended in health care settings.

Despite these observations and recommendations, relatively little is known about immunity to SARS-CoV-2 at the ocular surface, even though this may be an important route of transmission. Recent studies have documented SARS-CoV-2 specific antibody responses in tears following SARS-CoV-2 infection [4, 5], although the specificity, durability and functional significance of the antibodies detected after infection remains unclear.

Vaccination is highly protective from SARS-COV-2 infection and considerable evidence points towards neutralizing antibodies as a key correlate of protection [6]. Most neutralizing antibodies target the receptor binding domain (RBD) of the Spike protein which is critical in binding to cellular receptors [7]. RBD-specific antibodies correlate strongly with functional neutralizing antibody assays [8]. Little is known about whether vaccination against SARS-CoV-2 via the intramuscular route induces SARS-COV-2 antibodies in tears, especially RBD-specific antibodies. Herein, we collected basal tears, saliva and plasma samples from subjects with prior COVID-19 infection or vaccination and analyzed IgG and IgA antibody responses to a range of SARS-CoV-2 antigens.

## Methods

We enrolled participants (i) with and without prior SARS-CoV-2 infection from a previously described cohort [9] and (ii) prior to and following the Comirnaty (Pfizer-BioNTech) vaccine to donate blood, saliva and tears (Table 1). Basal (non-stimulated) tear samples (∼7μl/eye) were collected by capillary flow (Drummond) from the inferior tear meniscus as previously reported [10]. Saliva was collected by SalivaBio Oral Swabs (Salimetrics) following manufacturer’s instructions.

**Table 1:** Demographics and clinical characteristics of healthy individuals, COVID-19 patients and Comirnaty (Pfizer-BioNtech) vaccine recipients.

| Variables | Healthy Controls<br>(n=11) | COVID-19 Patients<br>(n=16) | Vaccine Recipients<br>(n=15) |
| --- | --- | --- | --- |
| Age, mean (range), years | 46.0 (21 – 64) | 52.7 (22 – 76) | 34.0 (25 – 57) |
| Gender |  |  |  |
| Female | 3 (27.3%) | 8 (50.0%) | 10 (66.7%) |
| Male | 8 (72.7%) | 8 (50.0%) | 5 (33.3%) |
| Time from symptom onset till sample collection, mean (range), days |  | 210.4 (65 – 249) |  |
| Severity |  |  |  |
| Mild |  | 6 (37.5%) |  |
| Moderate |  | 8 (50.5%) |  |
| Severe |  | 2 (12.5%) |  |
| Received Comirnaty (Pfizer-BioNtech) vaccine |  |  | 15 (100.0%) |
| Time after 1<sup>st</sup> vaccine to sample collection, mean (range), days |  |  | 37.3 (33 – 47) |
| Time after 2<sup>nd</sup> vaccine to sample collection, mean (range), days |  |  | 13.4 (12 – 18) |

SARS-CoV-2 specific IgG, IgA1 and IgA2 antibodies in plasma (1:200), saliva (1:5 and 1:50) and tears (1:5 and 1:50) from the respective cohorts (i and ii) were assessed by a customized multiplex bead array consisting of 5 SARS-CoV-2 proteins, including a whole Spike trimer (ST), the Spike 1 (S1), Spike 2 (S2), Receptor Binding Domain (RBD) of Spike and NP (Nucleoprotein) as previously described [11]. The SIVgp120 protein (Sino Biological) and uncoupled BSA-blocked beads were included as negative controls for background subtraction [11].

All participants provided written informed consent; the study was approved by the University of Melbourne human research and ethics committee (2056689 and 21198153983). Statistical analysis was performed with GraphPad Prism 9 (GraphPad Software). Antibody levels between antigens (ST, S1, S2, RBD, NP) within each sample type (plasma, tears, saliva) were compared using Kruskal-Wallis test (i) or Friedman test (ii) respectively followed by Dunn’s test for multiple comparisons.

## Results

42 participants were enrolled: 11 healthy controls, 16 with prior SARS-CoV-2 infection with a mean of 210 days from previous symptom onset, and 15 Comirnaty vaccine recipients, where samples were taken prior to vaccination and a mean of 13 days following their second vaccine dose (Table 1).

IgG antibodies to Spike proteins ST, S1, and S2 and to the NP protein were detected in tears, saliva and plasma from subjects with prior SARS-CoV-2 infection in comparison to uninfected controls. While RBD-specific antibodies were detected in convalescent plasma, RBD-specific antibodies were not detectable in convalescent tears and saliva in comparison to healthy controls (Figure 1a).

**Figure 1:**
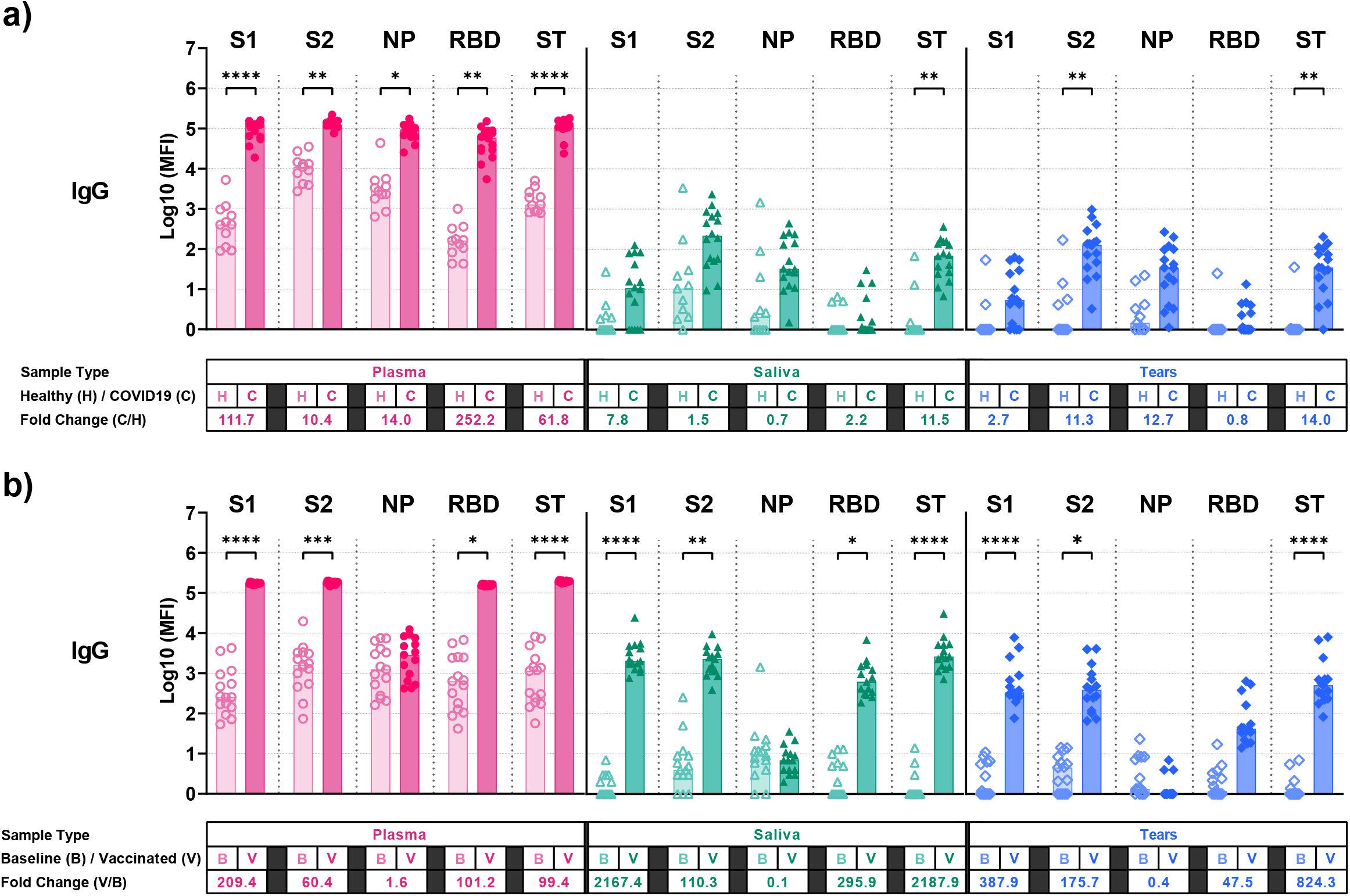
Anti-SARS-CoV-2 IgG found in plasma, saliva and tears of (a) convalescent COVID-19 patients and (b) Comirnaty (Pfizer-BioNTech) vaccinees. The presence of IgG specific for SARS-CoV-2 spike 1 (S1), spike 2 (S2), nucleoprotein (NP), receptor-binding domain (RBD) and whole spike trimer (ST) was compared in samples from (a) both healthy individuals (H) and convalescent COVID-19 patients (C; Kruskal-Wallis test), (b) as well as in paired baseline pre-vaccination (B) and 2 weeks post-second dose vaccinated (V) samples from Comirnaty (Pfizer-BioNTech) vaccinees (Friedman Test). MFI readings from tears and saliva samples were normalized to a final dilution of 1:200. Fold changes were calculated between grouped samples for the same antigen. *p-value: * < 0*.*05; **<0*.*01; ***<0*.*001; ****<0*.*0001*.

The Cormirnaty vaccine is highly protective for SARS-CoV-2 infection [12]. We found vaccination induced high levels of Spike-specific IgG antibodies (including to RBD) in tears, saliva and plasma (Figure 1b). As expected, IgG responses to NP were minimal, since the Comirnaty vaccine does not express this antigen.

IgA antibodies at mucosal surfaces are important in protection from multiple infectious diseases [13] and we previously observed IgA1 and IgA2 antibodies to Spike in the plasma of subjects for up to 4 months after SARS-CoV-2 infection [14]. We studied IgA1 and IgA2 antibodies in the plasma, tears, and saliva samples of subjects a mean of 7 months after SARS-CoV-2 infection (Figure 2a-b). Plasma IgA1 and IgA2 responses were detected to most Spike SARS-CoV-2 antigens in plasma but uniformly at lower levels than IgG antibodies. Plasma IgA1 and IgA2 responses were not detectable to NP 7 month following infection. No IgA1 and IgA2 responses to SARS-CoV-2 antigens were detected in tears and saliva. Background levels of cross-reactive IgA1 and IgA2 antibodies in uninfected controls were uniformly higher than for IgG responses.

**Figure 2:**
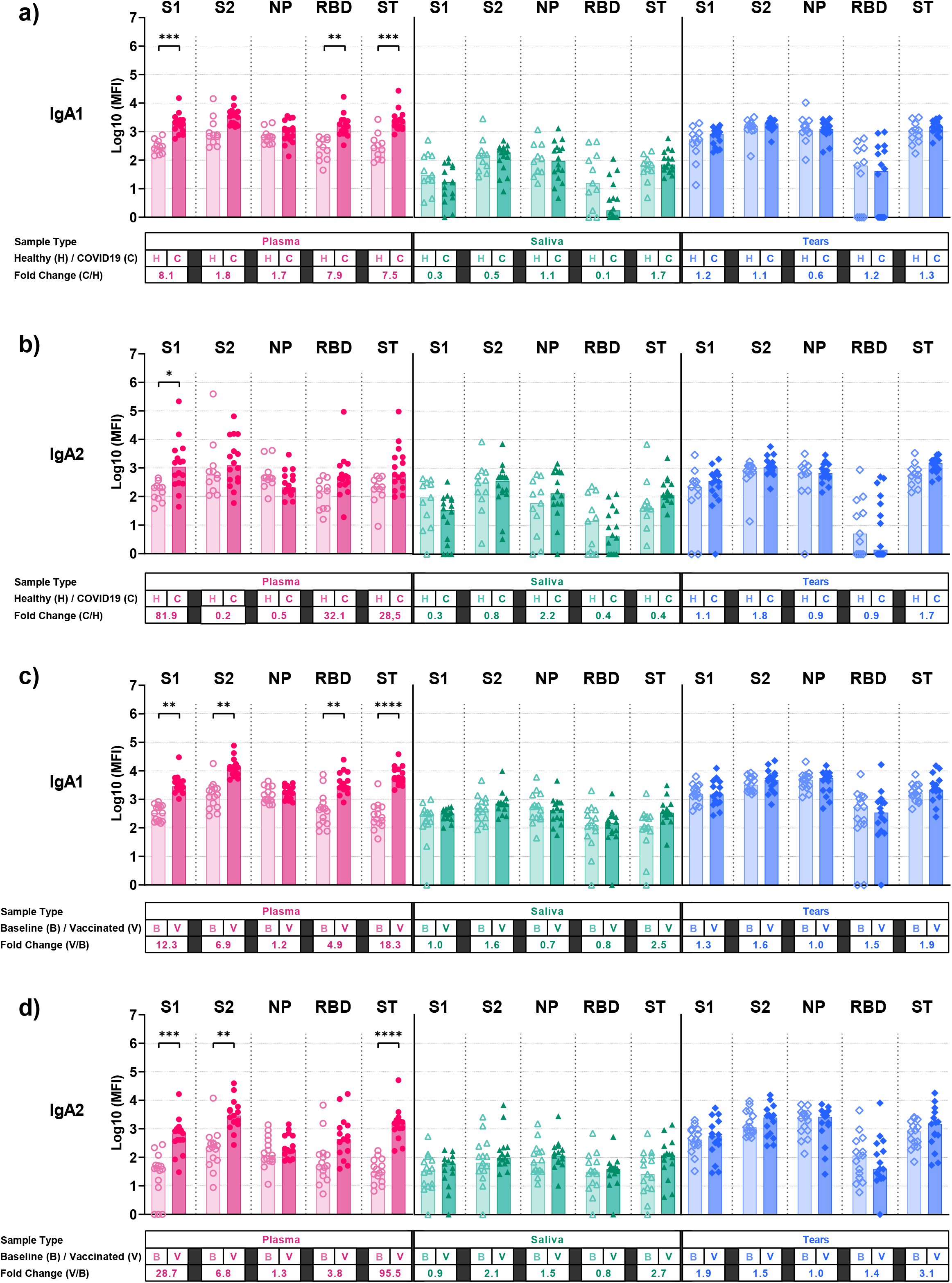
Anti-SARS-CoV-2 IgA1 and IgA2 found in plasma, saliva and tears of (a-b) convalescent COVID-19 patients and (c-d) Comirnaty (Pfizer-BioNTech) vaccinees. The presence of IgA1 and IgA2 specific for SARS-CoV-2 spike 1 (S1), spike 2 (S2), nucleoprotein (NP), receptor-binding domain (RBD) and whole spike trimer (ST) was compared in samples from (a-b) both healthy individuals (H) and convalescent COVID-19 patients (C; Kruskal-Wallis test), (c-d) as well as in paired baseline pre-vaccination (B) and 2 weeks post-second dose vaccinated (V) samples from Comirnaty (Pfizer-BioNTech) vaccinees (Friedman Test). MFI readings from tears and saliva samples were normalized to a final dilution of 1:200. Fold changes were calculated between grouped samples for the same antigen. *p-value: * < 0*.*05; **<0*.*01; ***<0*.*001; ****<0*.*0001*.

Intramuscular vaccination is not considered an optimal vaccination route to induce IgA responses at mucosal surfaces although some IgA responses can be induced by this vaccination method [15, 16]. Plasma IgA1 and IgA2 responses were detected to all 5 SARS-CoV-2 antigens early after Comirnaty vaccination but uniformly at lower levels than IgG antibodies (Fig 2c-d). Although non-significant increases of IgA1 and IgA2 responses to spike trimer were detected in saliva (fold change 2.5 and 2.7 respectively) and tears (fold change 1.9 and 3.1 respectively) after vaccination, overall, IgA responses to spike subunit proteins in the mucosal samples were not detectable. Background pre-vaccination levels of IgA1 and IgA2 antibodies were again uniformly higher than for IgG responses.

## Discussion

Previous work has identified SARS-CoV-2 specific antibodies in the tears of subjects following COVID-19, although little is known about the specificity or longevity of tear antibodies [4], and nothing has been reported on tear antibodies following COVID-19 vaccination to our knowledge. We found that although SARS-CoV-2 antibodies are detectable in the tears and saliva of subjects with prior SARS-CoV-2 infection, levels of antibodies to RBD were low or undetectable by 7 months following infection. Since most neutralizing antibodies bind to the RBD portion of Spike, this suggests neutralizing antibodies in these mucosal samples are low. Additional studies of the direct virus neutralization capacity of tear antibodies will be helpful, although most assays require larger sample volumes that are difficult to obtain with tears. Re-infection with SARS-CoV-2 is being increasingly reported and we speculate that low mucosal antibodies may leave subjects with prior SARS-CoV-2 infection vulnerable to re-infection through these sites, particularly through eye surfaces.

Importantly, this report demonstrated vaccination induced high levels of tear and saliva IgG antibodies, including to RBD. This suggests vaccination, at least early after the course is completed, induces key antibodies at the ocular surface which may help protect from acquisition of SARS-CoV-2. The relative levels and durability of tear antibodies induced by differing COVID-19 vaccines remains to be determined. This may be an important factor governing the waning of vaccine-induced immunity and the need for future booster vaccinations.

We found SARS-CoV-2 specific IgA responses in tears and saliva were low or non-detectable either 7 months following infection or early following vaccination. As a mucosal infection, one might expect IgA responses in tears and saliva following infection. The lack of IgA in tears and saliva following infection may reflect the waning of immunity. We have previously reported IgA responses wane rapidly in plasma post infection[14]. Similarly, strong plasma IgA responses were induced after a single vaccine dose of Phase Clinical trials I of a protein-MF59 adjuvanted spike glycoprotein clamp, however rapidly waned despite a second dose [17]. Further studies of IgA responses in tears earlier following infection are warranted. The lack of IgA responses in tears and saliva following intramuscular Comirnaty vaccination may reflect that this route is relatively poor at inducing mucosal IgA responses [15]. Although this vaccine is highly protective, whether modified regimens that induce IgA responses at mucosal surfaces are more durably protective warrants further study. Further study of secretory IgA, more specific to mucosal IgA, is also warranted. In general, we also noted that background IgA1 and IgA2 responses to SARS-CoV-2 antigens were high in the tears and saliva of uninfected and unvaccinated subjects. It is possible that prior exposure to other human coronaviruses may induce high levels cross reactive IgA antibodies at baseline and studies of antibodies to other coronaviruses are warranted. More refined of methods to detect SARS-CoV-2 IgA antibodies in tears and saliva sre needed.

In conclusion, COVID-19 infection and Comirnaty vaccination induced SARS-CoV-2-specific IgG antibodies in tears and saliva. The detection of IgG antibodies to RBD in tears and saliva following vaccination suggests that neutralising antibodies are present at these sites and may have a protective role. Our findings reinforce the need for widespread vaccination and eye protection in settings where this is not yet possible.

## Data Availability

The source data underlying Figure 1–2 and Table 1 are provided as a source data file. All other data are available from the authors upon request.

## Acknowledgments

This work was supported by the Victorian government, the Medical Research Future Fund (MRFF) to JAJ, AWC, SJK and Emergent Ventures Fast Grant to AWC. MPD, JAJ, AWC and SJK are supported by NHMRC Fellowships

## Notes

### Competing Interest Statement

The authors have declared no competing interest.

### Author Declarations

All participants provided written informed consent; the study was approved by the University of Melbourne human research and ethics committee (2056689 and 21198153983).

